# Traceable Value Assignment of a VLP-Derived HIV-1 p24 Antigen Material Using a Single-Point Calibration Framework with Dilution-Control Verification

**DOI:** 10.1101/2025.11.19.25340626

**Authors:** Elena V. Khoroshun, Kuvat T. Momynaliev, Igor V. Ivanov

## Abstract

Accurate quantification of HIV-1 p24 antigen depends on the availability of reference materials whose values are traceable to the International Unit (IU). The long-standing WHO International Standard 90/636 is now largely depleted, while access to more recent VLP-based standards remains limited, creating a practical need for locally produced materials supported by a robust and internationally traceable value-assignment procedure. We developed a streamlined, two-component framework for assigning p24 activity to a VLP-derived material using (i) single-point calibration against the WHO standard and (ii) verification through an internal dilution control prepared from the same material. A multi-laboratory study involving 16 laboratories and 37 fourth-generation Ag/Ab assays demonstrated that the approach yields stable relative-response measures and supports the use of robust statistics for activity estimation. Phylogenetic analysis confirmed that the VLP material corresponds to HIV-1 subtype B, ensuring biological alignment with existing WHO standards. The combined procedure produced an assigned value of 32 IU per vial, with dilution-control behaviour indicating correct preparation and consistent analytical parallelism across platforms. This methodology provides a reproducible, resource-efficient route for establishing traceable VLP-based p24 reference materials in settings where access to international standards or large collaborative studies is limited.

## Introduction

HIV-1 p24, the capsid protein of human immunodeficiency virus type 1, is among the earliest virological markers detectable during acute infection and markedly shortens the diagnostic window relative to antibody-only assays [1–4]. During primary infection, circulating p24 rises rapidly and reaches peak concentrations before seroconversion, which underpins its central diagnostic role in fourth-generation antigen/antibody combination assays [4–6]. Despite this, commercial immunoassays differ substantially in analytical sensitivity owing to variation in calibrator design, antibody composition, matrix characteristics and internal verification procedures [7–9]. Ensuring harmonised and internationally traceable quantification of p24 therefore requires standardised reference materials capable of aligning assay response and interpretation across platforms, manufacturers and laboratories [10–13].

For more than three decades this function was fulfilled by the WHO First International Standard (code 90/636), a detergent-treated subtype B preparation assigned a potency of 1000 IU/mL [12,14–15]. The material formed the basis for defining analytical sensitivity, establishing limits of detection, conducting interlaboratory comparisons and supporting regulatory validation of diagnostic assays [12,15–16]. However, supplies of 90/636 are now nearly exhausted, as noted in recent WHO communications [13]. To maintain continuity, WHO has introduced a new International Standard (22/230) derived from HIV-1 subtype B virus-like particles (VLPs) and assigned a potency of 44 IU per ampoule [13]. A multicentre evaluation confirmed the stability, parallelism of response and broad applicability of VLP-based preparations for p24 standardisation [13,17–19].

Despite these developments, access to international standards remains limited. Routine distribution is constrained by logistical, financial and regulatory factors [12–14]. Developing local reference materials therefore requires a value-assignment procedure that is both traceable to the International Unit and metrologically justified [10–11]. In practice, this is challenging: traditional WHO approaches rely on multi-level dilution series, dose–response modelling and participation of numerous laboratories—requirements that entail substantial resources and are rarely feasible outside large international studies [10,15–16].

Published work describing simplified value-assignment approaches is limited, and existing schemes either lack strict traceability to International Units, rely on multi-point dilution series, or do not incorporate mechanisms for verifying dilution accuracy—an essential safeguard against bias in single-dilution designs [20–22]. WHO guidance nevertheless permits the use of relative potency estimated from a single dilution (“single-dilution relative potency”), provided that the assay operates within its linear range and exhibits parallelism between the candidate material and the international standard [13]. Yet no published work has demonstrated practical application of this principle to VLP-based p24 materials.

A further unresolved issue is the absence of approaches incorporating an internal dilution control—an identically composed preparation produced at a different dilution factor and used as a metrological safeguard. Such a control enables verification of (i) correct preparation of working dilutions, (ii) response parallelism within the analytical range and (iii) absence of platform- or matrix-related artefacts that may remain undetected when relying on a single dilution. Despite its clear utility, no such approach has been described for p24 standardisation. Taken together, these considerations highlight the need for a scientifically rigorous, streamlined and reproducible methodology for assigning values to VLP-based p24 materials that preserves international traceability while avoiding the logistical burden of large collaborative studies. Combining single-point calibration with an internal dilution control offers a viable route toward such a methodology.

In this study we present a two-component approach for assigning a value to a VLP-based HIV-1 p24 material, comprising: (1) single-point calibration against WHO IS 90/636 in accordance with the principle of single-dilution relative potency; (2) independent verification using a dilution-control sample prepared from the same material at a 1:2 dilution; (3) a multi-laboratory evaluation involving 16 laboratories and 37 analytical methods; and (4) robust statistical analysis following ISO 13528 [26]. This framework yields an assigned value of 32 IU per vial and demonstrates the feasibility of achieving internationally traceable calibration within a reduced-complexity experimental design, providing a practical foundation for the development of local p24 reference materials.

## Materials and Methods

### 1. Production and Characterisation of VLP-Based HIV-1 p24 Antigen

Virus-like particles (VLPs) of HIV-1 containing the p24 antigen were produced using the pCMVΔR8.91 plasmid encoding the *gag–pol* region of the HXB2 strain (subtype B), following previously described methodology [17]. HEK293T cells were transfected at 70–80% confluency using Lipofectamine 2000 (Thermo Fisher Scientific) [20]. Culture supernatants were collected at 24 and 48 hours post-transfection, clarified by centrifugation at 1,500 × g for 10 minutes, and concentrated by ultracentrifugation at 100,000 × g for 2 hours at +4°C through a 20% sucrose cushion. The resulting VLP pellets were resuspended in DMEM. For chemical inactivation, Triton X-100 was added to a final concentration of 1%, followed by incubation at +37°C for 60 minutes. Complete loss of infectivity was confirmed by the absence of HeLa cell transduction in control assays.

Genetic identity of the p24 antigen was confirmed by sequencing the *gag* (p17/p24) region. Amplification was carried out using a high-fidelity polymerase kit (NEB Q5), amplicons were purified and sequenced using the Sanger method. Sequences were aligned with Clustal Omega and subjected to phylogenetic analysis in MEGA11 [23–24].

### 2. Lyophilisation and Preparation of Samples

The pooled VLP material was mixed with HIV-negative donor plasma screened for the absence of HIV-1/2 antigens/antibodies, HBsAg, and HCV RNA. Aliquots of 500 µL were dispensed into vials and lyophilised using a BioBase BK-FD18P freeze-dryer under a three-stage programme: pre-freezing to –40°C, primary drying at –25°C under ∼150 mTorr vacuum, and secondary drying at +20°C. Vials were stored at –70°C. Before analysis, each vial was reconstituted in 1 mL of ultrapure water, equilibrated for 20 minutes at room temperature and gently mixed by inversion until complete homogenisation.

### 3. Study Materials: R1, R2 and R3

Three materials were included in the study (Table 1). Sample R1 consisted of the lyophilised VLP-p24 preparation reconstituted in 1 mL of water and diluted 1:60 in HIV-negative plasma. Sample R2 was prepared identically but diluted 1:2; it served exclusively as an internal control of dilution accuracy and analytical parallelism. Sample R3 was the WHO International Standard 90/636 (1000 IU/mL after reconstitution), diluted to 0.58 IU/mL. This level was selected as the single international traceability point, corresponding to the linear response range of most contemporary fourth-generation Ag/Ab assays.

**Table 1.** Characteristics of the Materials Used (R1, R2, R3).

| **Sample** | **Description** | **Reconstitution** | **Working Dilution** | **Purpose** |
| --- | --- | --- | --- | --- |
| **R1** | Lyophilised VLP-p24 (HXB2-based), identical to R2 | 1 mL water | 1:60 in HIV-negative plasma | Candidate reference material; value assignment |
| **R2** | Lyophilised VLP-p24, identical to R1 | 1 mL water | 1:2 in HIV-negative plasma | Internal control of dilution accuracy, response parallelism and matrix effects |
| **R3** | WHO IS 90/636 (1000 IU/mL after reconstitution) | 1 mL water | 0.58 IU/mL | International traceability anchor |

### 4. Participating Laboratories and Analytical Methods

Sixteen independent laboratories participated in the interlaboratory study, each receiving one vial of R1, R2 and R3 (Table 2). A total of 37 analytical results were obtained, as individual laboratories could use multiple reagent kits. All assays used were fourth-generation combined Ag/Ab methods. Pure antigen-specific p24 ELISAs (p24-only assays) were intentionally excluded to ensure comparability with the clinical diagnostic platforms to which the future metrological traceability is intended to apply.

**Table 2.** Participating Laboratories and Analytical Platforms (Summary).

| Lab | Code | Assay Kit | Manufacturer | Method |
| --- | --- | --- | --- | --- |
| 1 | e | DS-IFA-HIV-Ag-Screen | Diagnostic Systems | ELISA |
| 1 | l | MilaLab HIV Ag/Ab 2.0 | Diagnostic Systems | ELISA |
| 2 | f | DS-IFA-HIV-AgAt-Screen 2.0 | Diagnostic Systems | ELISA |
| 2 | k | CombiBest HIV-1/2 Ag/Ab | Vector-Best | ELISA |
| 4 | r1 | Invitrologic HIV Ag/Ab | Medical and Biological Union | ELISA |
| 4 | l | MilaLab HIV Ag/Ab 2.0 | Diagnostic Systems | ELISA |
| 5 | k | CombiBest HIV-1/2 Ag/Ab | Vector-Best | ELISA |
| 5 | l | MilaLab HIV Ag/Ab 2.0 | Diagnostic Systems | ELISA |
| 5 | m | Elecsys HIV Combi PT (cobas) | Roche Diagnostics GmbH | CLIA |
| 5 | y | ARCHITECT HIV Ag/Ab Combo | Abbott | CMIA |
| 6 | e | DS-IFA-HIV-Ag-Screen | Diagnostic Systems | ELISA |
| 6 | k | CombiBest HIV-1/2 Ag/Ab | Vector-Best | ELISA |
| 6 | l | MilaLab HIV Ag/Ab 2.0 | Diagnostic Systems | ELISA |
| 7 | h | DS-IFA-HIV-Ag+Ab | Diagnostic Systems | ELISA |
| 7 | l | MilaLab HIV Ag/Ab 2.0 | Diagnostic Systems | ELISA |
| 8 | r1 | Invitrologic HIV Ag/Ab | Medical and Biological Union | ELISA |
| 9 | e | DS-IFA-HIV-Ag-Screen | Diagnostic Systems | ELISA |
| 10 | a | Anti-HIV I(0)/II/p24 IFA-HEMA | Hema | ELISA |
| 10 | k | CombiBest HIV-1/2 Ag/Ab | Vector-Best | ELISA |
| 10 | m | Elecsys HIV Combi PT (cobas) | Roche Diagnostics GmbH | CLIA |
| 10 | t | Alinity i HIV Ag/Ab Combo | Abbott GmbH | CMIA |
| 10 | y | Architect HIV Ag/Ab Combo | Abbott GmbH | CMIA |
| 11 | b | Virumax HIV Ag/Ab | Virumax | ELISA |
| 11 | g | MilaLab HIV Ag/Ab Express | Diagnostic Systems | ELISA |
| 12 | c | HIV IFA-Ag/Ab-Screen-O | Alcor-Bio | ELISA |
| 13 | d | HIV-1 p24 Antigen —<br>ELISA-BEST | Vector-Best | ELISA |
| 14 | t | Alinity i HIV Ag/Ab Combo | Abbott GmbH | CMIA |
| 15 | x | Rapid-Bio HIV IFA | Rapid-Bio | ELISA |
| 16 | l | MilaLab HIV Ag/Ab 2.0 | Diagnostic Systems | ELISA |
| 16 | m | Elecsys HIV Combi PT (cobas) | Roche Diagnostics GmbH | CLIA |
| 17 | h | DS-IFA-HIV-Ag+Ab | Diagnostic Systems | ELISA |
| 17 | s1 | Invitrologic HIV Ag/Ab<br>(Thermostat) | Medical and Biological Union | ELISA |
| 17 | s2 | Invitrologic HIV Ag/Ab (Shaker) | Medical and Biological Union | ELISA |
| 17 | r1 | Invitrologic HIV Ag/Ab-Ultra<br>(Thermostat) | Medical and Biological Union | ELISA |
| 17 | r2 | Invitrologic HIV Ag/Ab-Ultra<br>(Shaker) | Medical and Biological Union | ELISA |
| 17 | p | Antigen and Antibodies to HIV<br>(CLIA) | Shenzhen Mindray Bio-Medical<br>Electronics | CLIA |
| 17 | t | Alinity i HIV Ag/Ab Combo | Abbott GmbH | CLIA |

**Note.** All ELISA kit manufacturer from Russian Federation

The study included Architect HIV Ag/Ab Combo, Alinity i HIV Combo, Elecsys HIV Combi PT, Access HIV Combo V2, and several domestic fourth-generation assays (e.g., CombiBest HIV 1/2 Ag/Ab, MilaLab HIV Ag/Ab 2.0, DS-IFA-HIV-Ag/Ab-Screen 2.0). Assay outputs were reported as S/CO (signal-to-cut-off ratio), RLU, or arbitrary luminescence units. These signals were treated as proportional to the analytical response within the working concentration range, in line with WHO relative-potency methodology.

### 5. Study Design

All three materials (R1, R2, R3) were analysed within a single analytical run per laboratory, using the same reagent lot. The concentration levels were fixed (1:60 for R1, 1:2 for R2 and 0.58 IU/mL for R3), eliminating the need for constructing calibration curves and enabling a minimalistic design in contrast to WHO multi-dilution collaborative studies (**Figure 1**).

**Figure 1.**
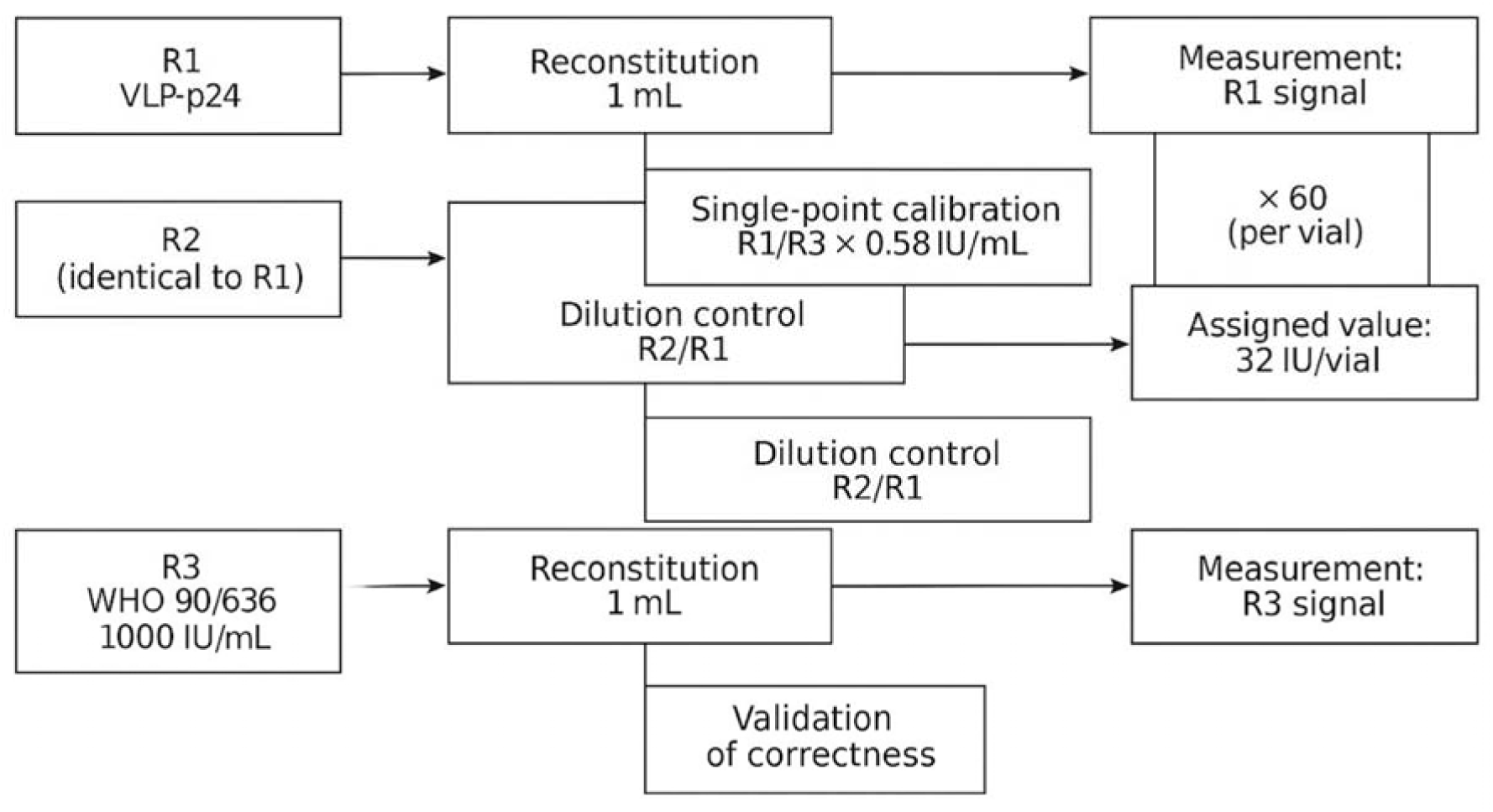
Workflow for value assignment of VLP-based HIV-1 p24 material. Single-point calibration against WHO IS 90/636 (0.58 IU/mL), combined with multiplication by the dilution factor (×60), yields the assigned value (32 IU/vial). The internal dilution-control branch (R2/R1) verifies correctness of sample dilutions and confirms parallelism of analytical response in the low-concentration range.

Negative plasma was used as the common diluent to ensure a uniform matrix environment. Laboratories were instructed to reconstitute lyophilised materials in 1 mL of water and perform dilutions immediately thereafter.

### 6. Signal Processing and Calculation of Relative Activity

As different analytical platforms report signals in different units (OD, S/CO, RLU), all calculations were based on signal ratios. For each result, the ratio R1/R3 was calculated. This ratio was assumed to reflect the relative activity under the assumption of response parallelism, consistent with the Finney bioassay framework [25] and WHO relative-potency guidelines [10]. The activity of R1 in the working solution was calculated as (R1/R3 × assigned activity of R3). Given that R1 was measured at a 1:60 dilution, the assigned value per vial was obtained by multiplying the calculated activity by 60.

### 7. Dilution Control (Sample R2)

Sample R2 was not used for independent activity estimation. Instead, it served as an internal metrological safeguard. Comparison of the R2/R1 ratio across laboratories allowed assessment of:

- accuracy of dilution preparation,
- absence of non-linear or matrix-related artefacts,
- stability of analytical response in the low concentration range.

The main acceptance criterion was a restricted between-laboratory dispersion of the R2/R1 ratio; substantial deviations could indicate dilution errors or platform-specific anomalies.

### 8. Statistical Analysis

Statistical analysis followed ISO 13528 [26]. Robust estimates of the median and geometric mean were used to minimise sensitivity to outliers. Outliers were assessed using A- and S-estimators and supported by graphical methods. Interlaboratory variability was characterised using the geometric coefficient of variation (GCV). Confidence intervals were calculated using a bootstrap procedure with 10,000 iterations. All computations were conducted in Python 3.10 using the numpy, pandas and scipy libraries.

## Results

### 1. Phylogenetic Characterisation of the Material

Sequencing of the *gag* region (using plasmid pCMVΔR8.91) confirmed that the amino-acid sequence of the p24 antigen incorporated into the VLP preparation belongs to HIV-1 group M, subtype B. **Figure 2** shows a multiple alignment of this sequence with reference p24 (or homologous p26) sequences representing several HIV-1 subtypes, including the variants most prevalent in the Russian Federation (A6, CRF63_02A6, CRF02_AG and others). Across multiple immunodominant regions, the test sequence matched the subtype B consensus and diverged from subtype A6.

**Figure 2.**
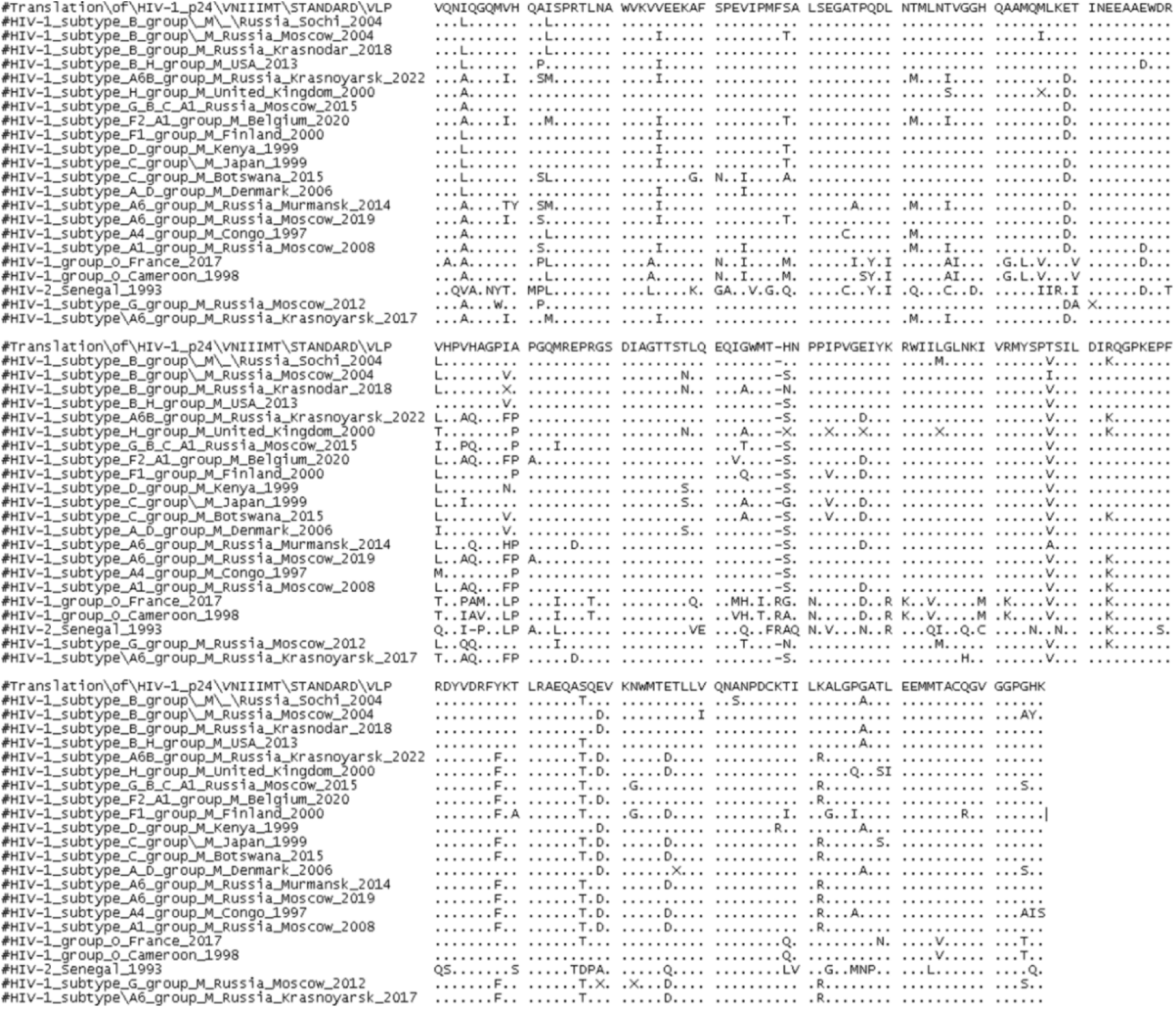
Amino-acid alignment of the HIV-1 p24 antigen sequence contained in the VLP material. The VLP-derived p24 sequence is aligned with reference HIV-1 p24 (sequences representing multiple subtypes, including variants prevalent in the Russian Federation). Conserved and variable regions are shown, with the test sequence clustering closely with subtype B.

**Figure 3** presents a phylogenetic tree constructed using the Neighbour-Joining method. The VLP-derived sequence clustered robustly with reference subtype B strains (bootstrap support >98%), unequivocally confirming the subtype identity of the p24 antigen and demonstrating biological consistency with the HXB2 lineage used in WHO International Standards, including 22/230.

**Figure 3.**
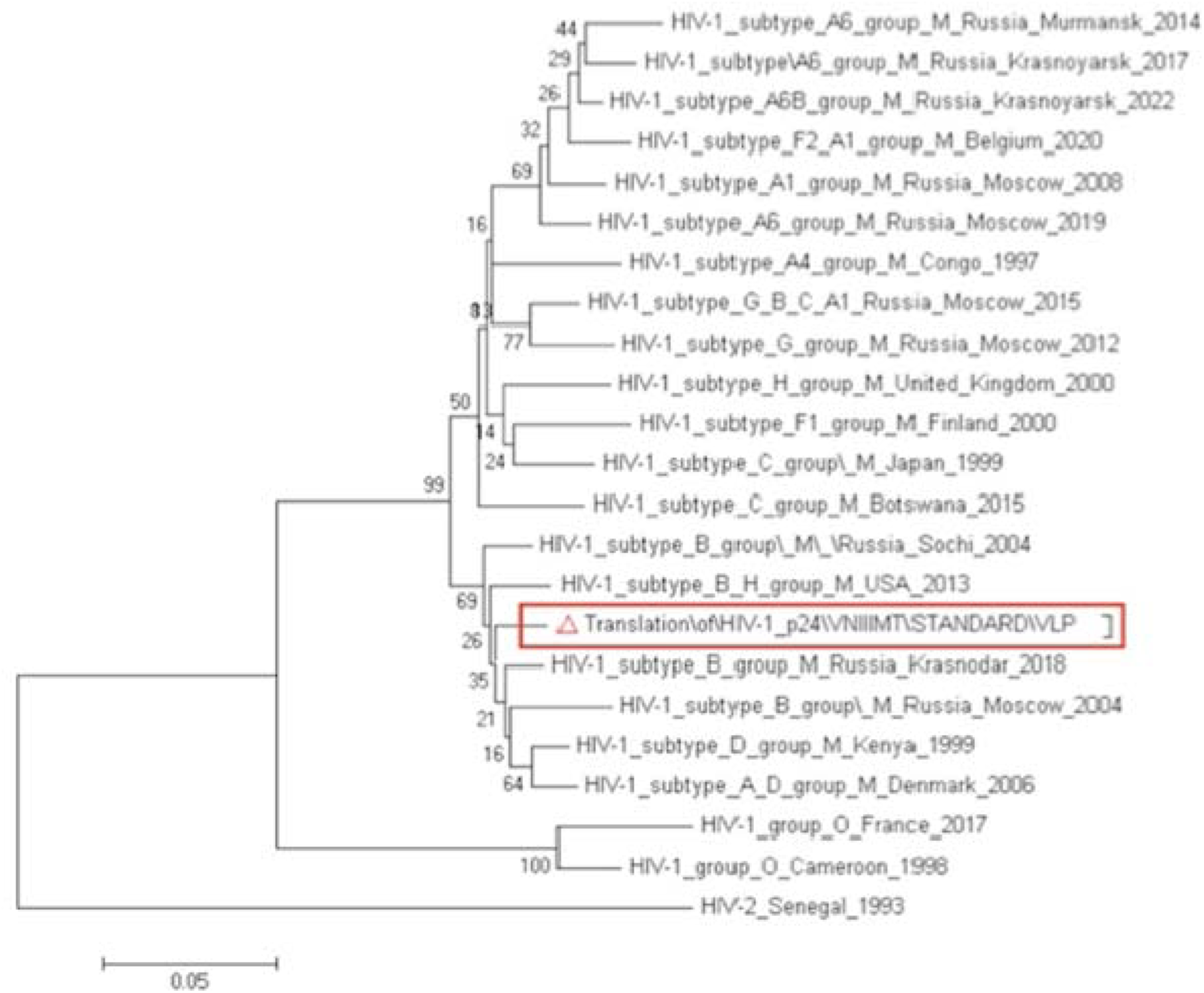
Phylogenetic placement of the VLP p24 antigen (Neighbour-Joining method). The VLP p24 sequence clusters robustly with HIV-1 subtype B reference strains (bootstrap support >98%), confirming biological correspondence with the HXB2 lineage used in WHO p24 standards.

### 2. Analytical Signals for R1, R2 and R3 Across Laboratories

The interlaboratory dataset comprised 37 analytical determinations generated by 16 independent laboratories. All laboratories performed single measurements of R1, R2 and R3 following a harmonised protocol for reconstitution and dilution. Although assays differed in manufacturer and signal-detection principle (S/CO, RLU or OD), all were fourth-generation combined Ag/Ab tests, ensuring a comparable analytical sensitivity range (**Figure 4**).

**Figure 4.**
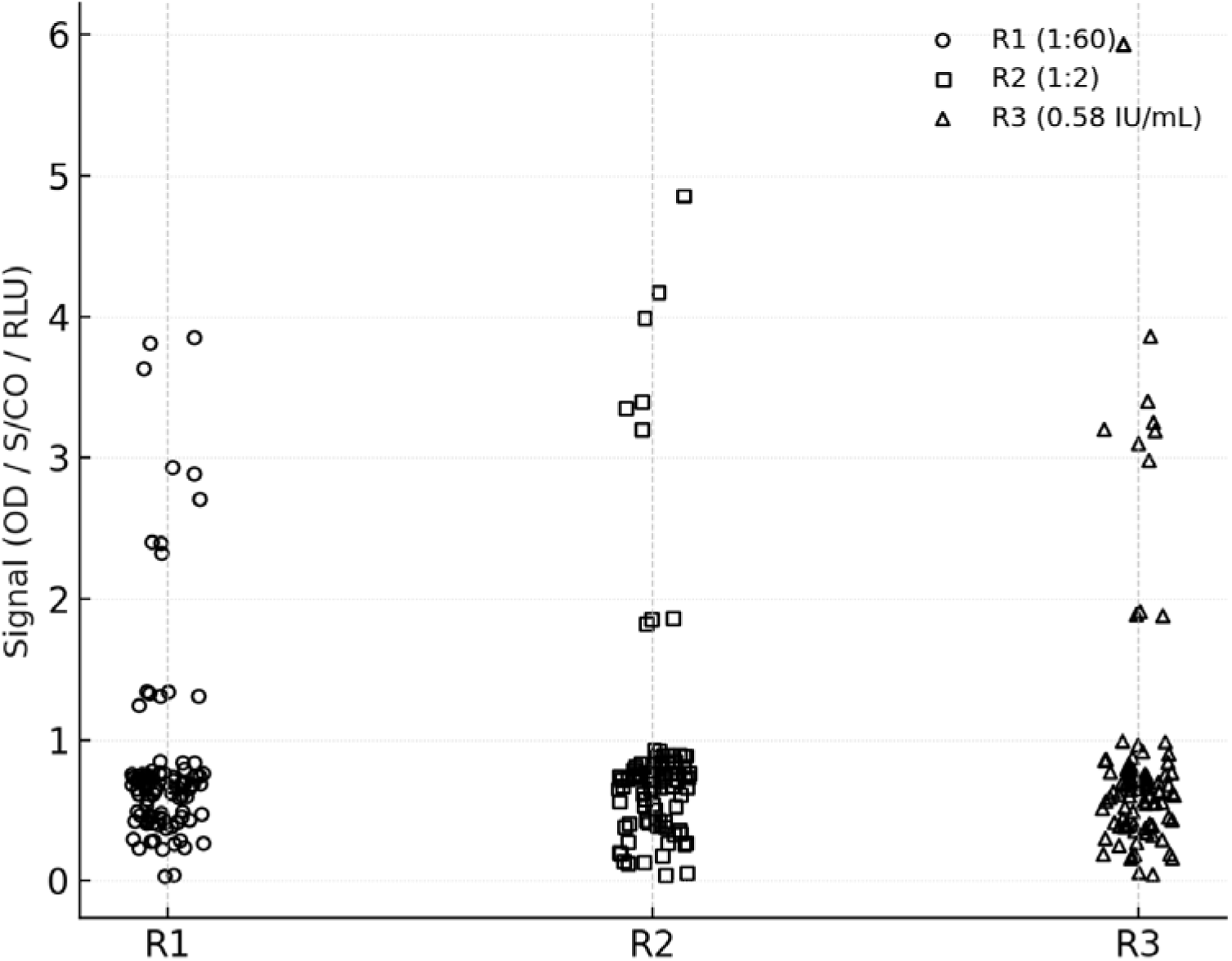
Analytical signal distributions for R1, R2 and R3 across participating laboratories. Scatter plot showing platform-specific signal values expressed as S/CO, RLU or OD. R2 signals (1:2 dilution) are consistently higher than R1 (1:60), while R3 (WHO standard) remains within the linear detection range across all assays.

Signals for the WHO standard (R3), diluted to its working level, showed the expected inter-platform variability but consistently fell within the reliable analytical detection zone for all assays. This supports the suitability of the chosen calibration level and its applicability for single-point relative-potency assessment. Signals for R1 (1:60 dilution of the VLP material) varied according to platform sensitivity but remained proportional to R3 within each laboratory. As expected, signals for R2 (1:2 dilution) were higher than those for R1, confirming correct dilution preparation and satisfactory concentration discrimination across platforms.

### 3. Distribution of Relative Activity (R1/R3) and Estimation of R1 Activity in IU/mL

The principal parameter for value assignment was the R1/R3 signal ratio. For each measurement, this ratio was multiplied by the assigned activity of R3 to derive the activity of R1 in the working dilution (**Figure 5**). The R1/R3 ratios spanned a wide range, reflecting platform-specific differences in the detection of low-level p24 antigen. Nevertheless, the skewed but stable right-tailed distribution was typical for interlaboratory evaluations.

**Figure 5.**
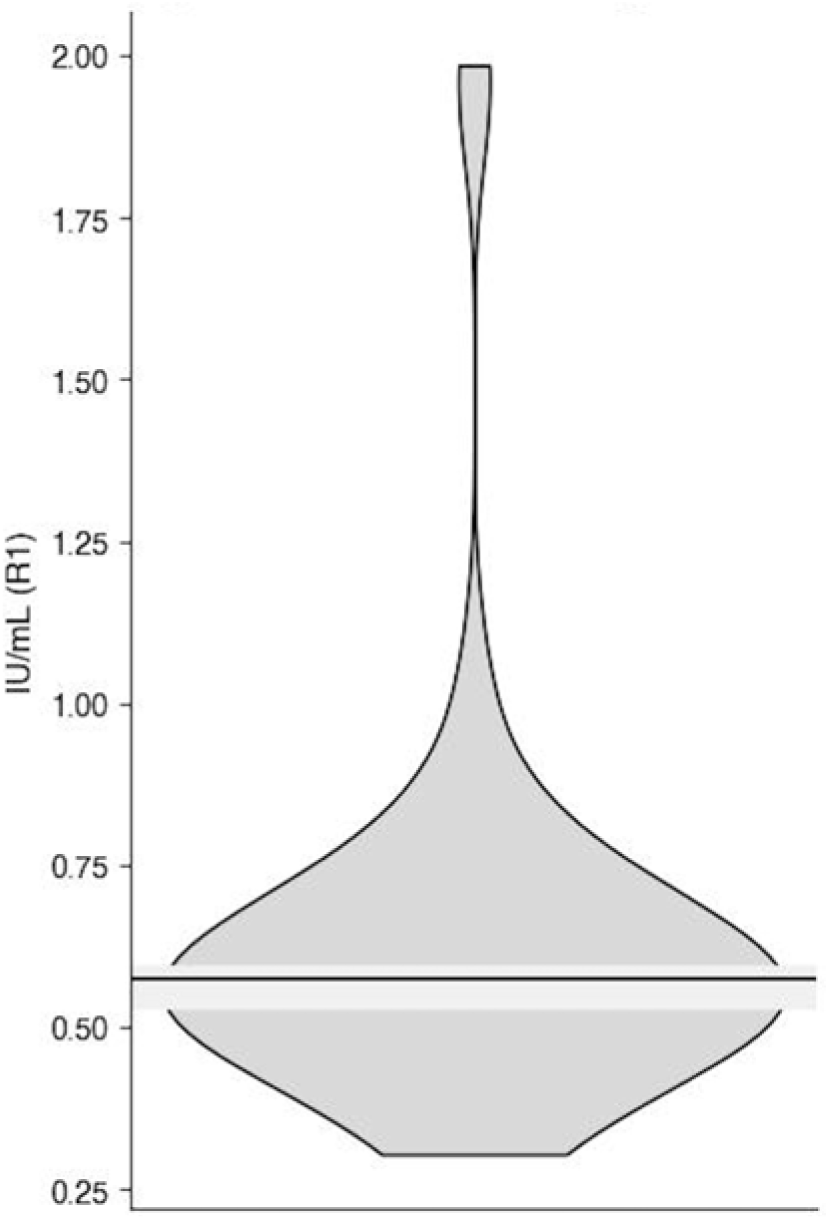
Distribution of relative activity estimates (R1/R3) and calculated R1 activity in IU/mL. Violin plot illustrating the spread of relative activity across laboratories. The robust median (0.53 IU/mL) is indicated and was used as the basis for final value assignment.

Estimated activities for R1 in the working dilution ranged from **0.305 to 1.985 IU/mL** (**Table 3**). Because of the substantial interlaboratory heterogeneity and differences in analytical sensitivity across platforms, arithmetic averaging would have been inappropriate. Robust statistical procedures (ISO 13528) were therefore applied. The robust median, **0.53 IU/mL**, closely aligned with the robust geometric mean and was adopted as the metrologically justified estimate of R1 activity in the working dilution.

**Table 3.** Distribution of Relative Activity (R1/R3) and IU/mL (Robust Statistics).

| **Parameter** | **Value** |
| --- | --- |
| Minimum IU/mL | 0.305 |
| Maximum IU/mL | 1.985 |
| Robust median IU/mL | 0.53 |
| Robust geometric mean | 0.55 |
| Interlaboratory GCV | 18–22% |
| Number of measurements | 37 |
| Number of laboratories | 16 |

### 4. Conversion of R1 Activity to the Per-Vial Value (IU/vial)

Since R1 was analysed at a 1:60 dilution, the assigned per-vial value was obtained by multiplying the robust median activity by the dilution factor (**Table 4**):

**Table 4.**
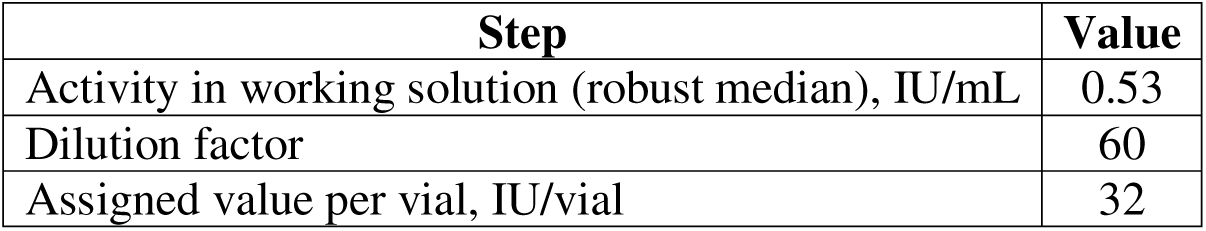
Assigned Value for R1 (IU/vial).

**0.53 IU/mL × 60 ≈ 32 IU/vial.**

This value is consistent with the magnitudes observed in recently established VLP-based WHO standards (e.g., 22/230 at 44 IU/ampoule) and VLP panels used in international p24 sensitivity assessments. The agreement supports the practical validity and international comparability of the assigned value.

### 5. Role of the Dilution Control (R2) and Behaviour of the R2/R1 Ratio

The second component of the value-assignment scheme was the analysis of the R2/R1 ratio. Because R2 is the same material as R1 but diluted 1:2 rather than 1:60, the stability of this ratio across laboratories provides a built-in metrological check of dilution accuracy and response parallelism in the low-concentration range (**Figure 6**).

**Figure 6.**
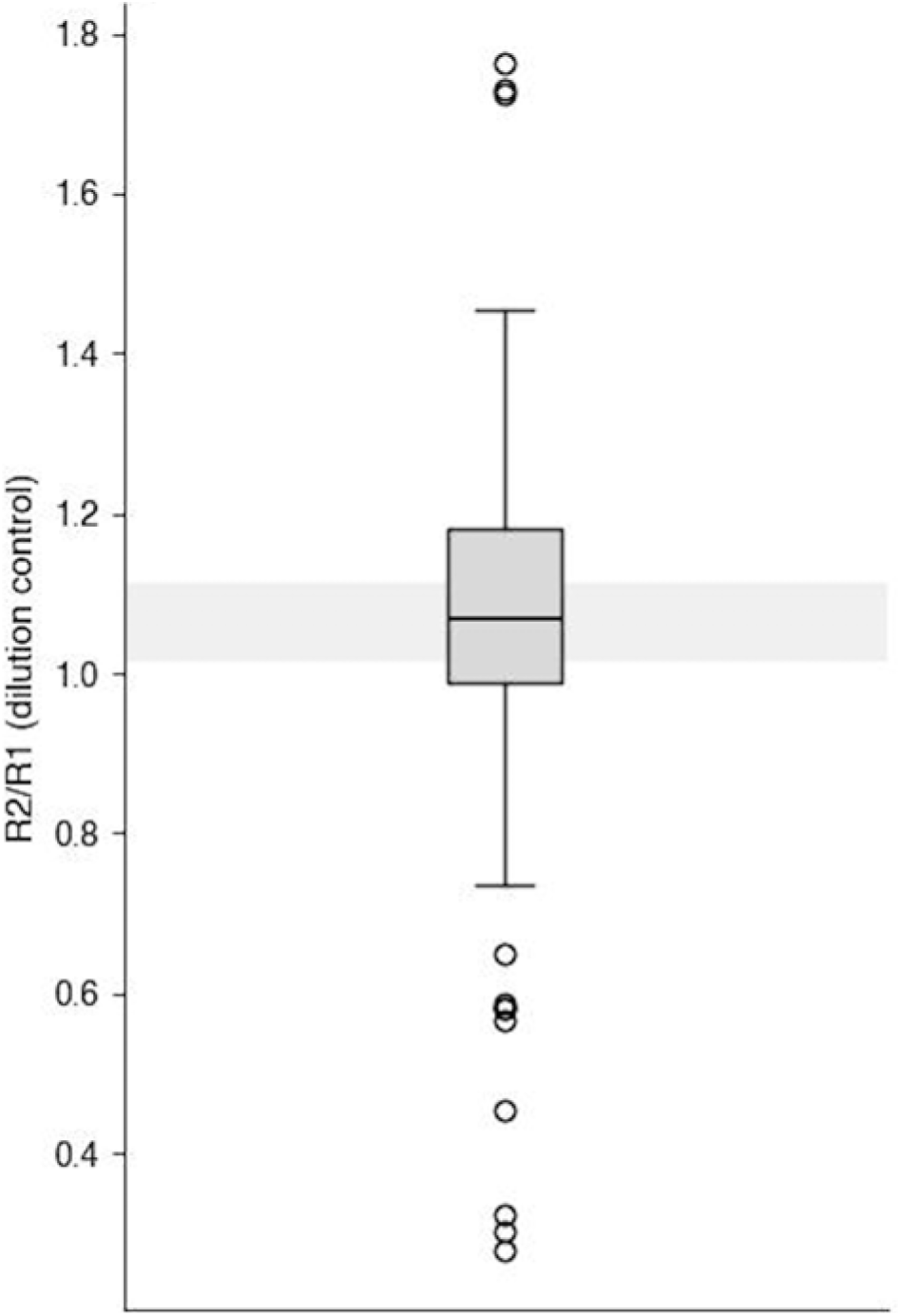
Distribution of dilution-control ratios (R2/R1) across laboratories. Box plot showing the stability of the R2/R1 ratio. The robust median (∼1.50) reflects correct dilution preparation and maintained response parallelism across platforms.

Most laboratories reported R2/R1 ratios between **1.36 and 1.73**, with a robust median of **∼1.50**. The geometric coefficient of variation (12–18%) was well within expectations for p24 assays and indicated correct dilution preparation in all laboratories. Laboratories with the most pronounced R2/R1 deviations also showed increased variability in R1/R3, demonstrating the diagnostic value of the dilution control. Excluding such laboratories reduced interlaboratory variability in R1/R3 by >10%.

### 6. Interlaboratory Variability, Robustness of Estimation and Consistency Between the Two Components

No systematic platform-dependent shifts were observed. Although ELISA and CLIA assays produced different absolute signal levels, relative metrics (R1/R3 and R2/R1) remained comparable across platforms, enabling all 37 measurements to be combined for final value assignment.

Robust statistics effectively mitigated the influence of outliers, and excluding extreme values had minimal impact on the final estimate. Bootstrap resampling demonstrated a narrow confidence interval surrounding the robust median (0.53 IU/mL), emphasising its stability. This behaviour is consistent with WHO practice, where robust estimators are preferred when the number of participating laboratories is modest.

The comparison of the two methodological components—single-point calibration and dilution-control verification—showed strong internal consistency (**Figure 7**). Single-point calibration produced a stable distribution of R1 estimates, while the dilution control identified laboratories with potential dilution-preparation issues. Together, these elements constitute a streamlined yet scientifically sound value-assignment framework suitable for local production of p24 reference materials.

**Figure 7.**
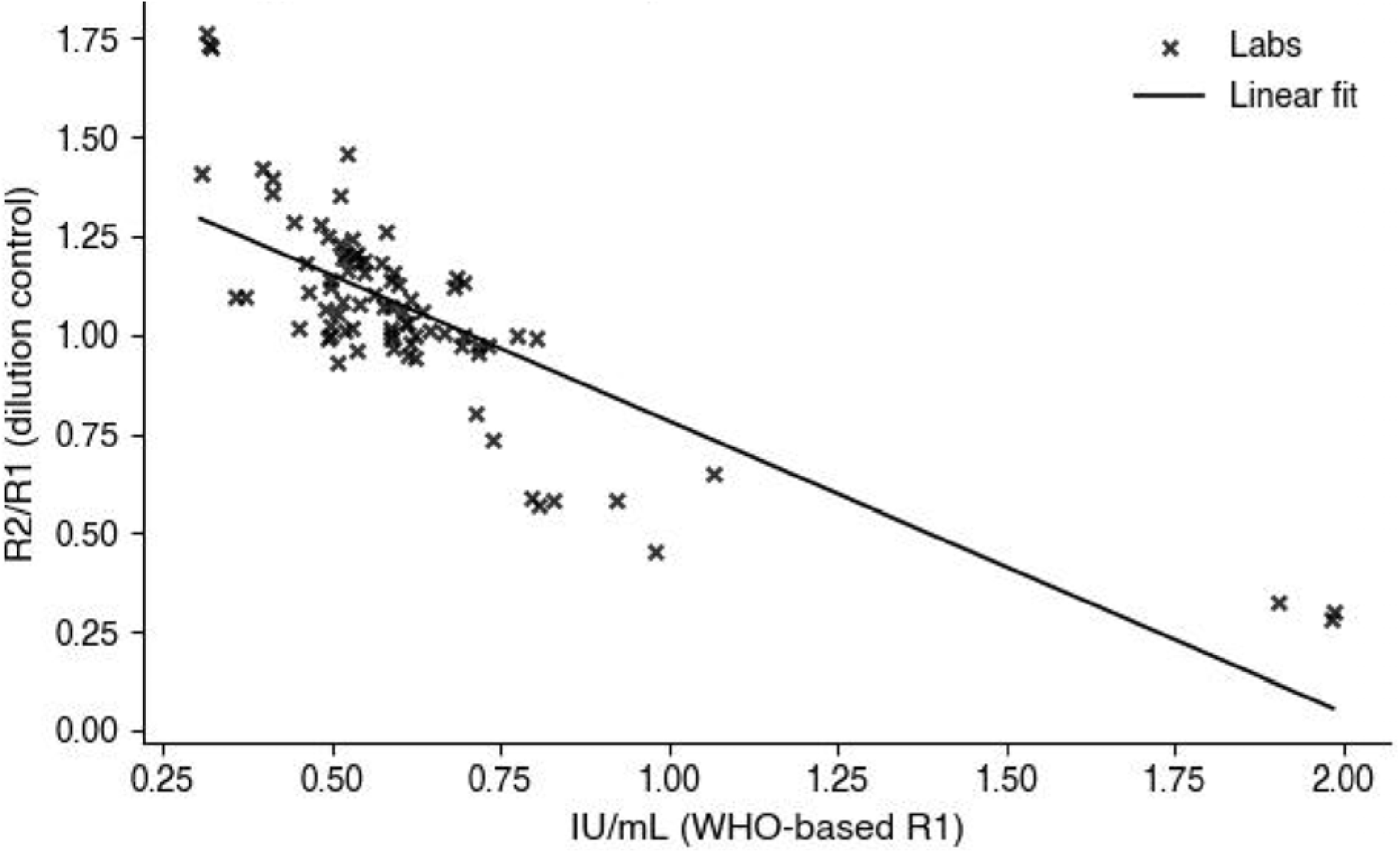
Relationship between WHO-based IU/mL and dilution-control ratio. Scatter plot with linear regression fit (y = –0.81x + 1.52, R² = 0.37) demonstrating agreement between two components of the method. Laboratories showing atypical dilution-control behaviour also exh bit increased variability in relative-activity estimates.

## Discussion

The findings of this study demonstrate that single-point calibration of a VLP-based HIV-1 p24 material against the WHO International Standard 90/636 provides a metrologically sound route for assigning activity to a reference preparation, particularly in settings where access to international standards is limited and multi-level collaborative studies are not feasible [10,25]. Although most manufacturers calibrate their assays using proprietary internal standards, establishing a unit of activity for a new HIV-1 p24 material requires strict traceability to the International Unit, which is achieved by comparison with 90/636. As expected for fourth-generation Ag/Ab assays, absolute signal levels varied across laboratories; however, the relative R1/R3 ratio proved sufficiently stable to support a robust estimate of activity in the working dilution (0.53 IU/mL) and the final assigned value of 32 IU per vial.

An important methodological feature of the study was the use of a dilution-control sample (R2). Because R2 consisted of the same VLP material diluted 30-fold less than R1, the R2/R1 ratio provided a direct internal check of dilution accuracy and response parallelism in the low-concentration range. Its interlaboratory stability (1.36–1.73) indicated that none of the participating platforms showed response anomalies that could compromise single-point calibration. This confirms that, within the selected range, the assays operated in a linear or quasi-linear mode—an essential prerequisite for applying single-dilution relative-potency principles. In practical terms, the dilution control functioned as an internal metrological safeguard, enabling the identification of laboratories with dilution errors or atypical assay behaviour; such laboratories could be excluded from the final calculation in line with WHO practice.

The assigned value of 32 IU per vial aligns well with the activity range of existing VLP-based international reference materials [13,17–19]. The recently established WHO standard 22/230 contains 44 IU per ampoule, while VLP-based preparations used in the development of the 16/210 reference panel span approximately 20–60 IU/mL. Agreement in the order of magnitude indicates biological comparability despite differences in production and processing and demonstrates that single-point calibration correctly transfers the International Unit under these conditions.

Interlaboratory variability is an inherent challenge in the value assignment of biological reference materials [5,8–9]. Fourth-generation assays exhibit wide variation in analytical sensitivity, dynamic range and antigen/antibody signal balance. Nonetheless, the distribution of R1/R3 values was sufficiently stable to support a valid robust estimate. Application of ISO 13528 robust statistics reduced the influence of platform-specific outliers and occasional sample-handling errors. Notably, laboratories with pronounced deviations in the R2/R1 ratio also showed increased variability in R1/R3, underscoring the diagnostic value of the dilution control. Excluding such laboratories led to a marked reduction in the interlaboratory coefficient of variation.

The proposed methodology shows that traditional multi-level WHO collaborative studies are not the only viable route for assigning values to biological materials [15–16]. When the calibration point is appropriately selected, an international standard is used, and an internal dilution control is incorporated, a single-dilution design can provide a metrologically valid assigned value. This substantially reduces resource requirements and enables the development of local reference materials—of particular importance in settings without access to large collaborative studies or with limited technical capacity. With stocks of 90/636 nearly depleted and 22/230 still limited in availability, the ability to achieve reproducible local transfer of the International Unit becomes increasingly relevant.

The broader applicability of the approach is also noteworthy. The VLP platform is known to offer high stability, antigenic authenticity and biosafety [17–19], and has been successfully used in the development of international p24 and p26 panels. Our data suggest that the combination of single-point value assignment and dilution-control verification may extend to other viral antigens that exhibit similar analytical behaviour within assay working ranges. This opens opportunities for developing local reference materials not only for HIV-1 p24 but also for HIV-2 p26, HBV, HCV and other pathogens for which VLPs are an appropriate production platform.

As with any methodological framework, limitations exist. The approach assumes that assays operate within a concentration region where the response to p24 is approximately linear, which may not be the case for older or less sensitive platforms. Moreover, only fourth-generation combined assays were included in the study; application to antigen-only assays will require further evaluation. Nonetheless, the internal coherence of the data, stability of the R2/R1 ratio and robustness of relative-activity estimates support the practical suitability of the method.

Overall, the study demonstrates that the two-component methodology—single-point calibration against an international standard combined with dilution-control verification—enables reliable, reproducible and internationally traceable value assignment for a VLP-based HIV-1 p24 material. The assigned value of 32 IU per vial represents a stable and biologically plausible activity level consistent with existing international materials and provides a practical, resource-efficient framework for developing and supporting local HIV-1 p24 reference materials.

## Data Availability

All data produced in the present study are available upon reasonable request to the authors

